# The optic radiations and reading development: a longitudinal study of children born term and preterm

**DOI:** 10.1101/2023.07.17.23292774

**Authors:** Lisa Bruckert, Garikoitz Lerma-Usabiaga, Lauren R. Borchers, Virginia A. Marchman, Katherine E. Travis, Heidi M. Feldman

**Author notes:** **Corresponding author:** Heidi M Feldman Address: Medical School Office Building, 1265 Welch Rd. Mail Code 5415, Stanford, CA 94305, USA.

## Abstract

**Purpose:** To determine if development of single-word reading between ages 6 and 8 years was related to change in a microstructural property of the optic radiations (OR), white matter circuits that are not typically associated with language or reading and if the patterns of association were similar in children born full term (FT) and preterm (PT).

**Methods:** FT (n=34) and PT (n=34) were assessed using the Woodcock Reading Mastery Test at 6, 7, and 8. Word Identification raw score was the outcome measure. Diffusion MRI was acquired at 6 and 8 years using a 96-direction scan (b=2500 sec/mm^2^). Probabilistic tractography identified left and right OR. We used linear mixed models to determine if change in fractional anisotropy (FA) averaged across the left and right OR was associated with growth in reading.

**Results:** The rate of reading growth was similar in both groups. FA of the OR was higher in children born FT than PT at age 8. Change in FA of the OR from age 6 to 8 was negatively associated growth in reading across both birth groups.

**Conclusion:** Individual differences in the rate of development in reading skills was associated with individual differences in the change in FA of the visual pathways in both children born FT and PT. Decreasing FA implicates increasing axonal diameter and/or complexity in fiber structure as the drivers of faster reading development.

**Highlights:**

- The rate of reading growth was similar in children born full term and preterm
- FA of the optic radiations increased from age 6 to 8 years in both groups
- FA of the optic radiations was higher in full term than preterm children at age 8
- Decreasing FA in optic radiations was associated with accelerated reading growth
- Associations of FA in optic radiations and reading were similar in two birth groups

## 1. Introduction

Children begin reading development around age 6 years, when they enter elementary school, learning to link letters to sounds and printed words to spoken words. By the time they reach around age 8 or 9, children can decode complex texts fluently and apply reading skills to learning about the world. The brain is undergoing development during this same period, including changes in the properties of white matter circuits essential to the transmission of information throughout the brain (Lebel et al. 2008; Lebel & Beaulieu 2011; Yeatman, Wandell & Mezer 2014). Many studies have found associations between early reading skills and properties of white matter pathways in the brain that are part of a language and reading network, including the arcuate fasciculus, superior longitudinal fasciculus, and inferior longitudinal fasciculus. (Beaulieu et al. 2005; Travis et al. 2016; Vandermosten et al. 2012; Yeatman et al. 2012). Properties of these circuits at young ages contribute to the prediction of reading skills at older ages (Borchers et al. 2018; Hoeft et al. 2011; Myers et al. 2014b; Wang et al. 2017). Associations of reading with other brain white matter circuits beyond the language network have not been thoroughly evaluated. Furthermore, few studies consider how the development of reading over time relates to changes in brain structure. The current study investigates change in a microstructural property of the optic radiations (OR), a pathway within the visual system, in relation to the development of reading between ages 6 and 8 years in a sample of children born at term and preterm.

Many neuroimaging studies using diffusion magnetic resonance imaging (dMRI) have shown that reading skills in typically developing children and adolescents correlate with contemporaneous diffusion metrics of language- and reading-related pathways (Beaulieu et al. 2005; Ben-Shachar, Dougherty, and Wandell 2007; Wandell and Yeatman 2013). Associations between children’s reading skills and diffusion metrics of the left arcuate fasciculus and left superior longitudinal fasciculus (Travis, Ben-Shachar, et al. 2016; Vandermosten et al. 2012; Yeatman et al. 2012) implicate the dorsal pathway in individual differences in reading (Hickok & Poeppel 2004; Scott & Wise 2004). Associations between children’s reading abilities and diffusion metrics of the left inferior longitudinal fasciculus and left inferior fronto-occipital fasciculus (Rollans et al. 2017; Yeatman et al. 2012) implicate the ventral pathway in individual differences in reading (Hickok & Poeppel 2004; Scott & Wise 2004). Differences between good and poor readers in white matter volume and diffusion metrics have been found during childhood, adolescence, and adulthood in these same pathways (Beaulieu et al. 2005; Darki et al. 2012; Deutsch et al. 2005; Klingberg et al. 2000; Niogi & McCandliss 2006).

Neuroimaging studies have also found that white matter pathways have proven useful in predicting later reading abilities. Diffusion metrics of reading-related pathways at younger ages have been found to predict reading skills at older ages (Borchers et al. 2018; Hoeft et al. 2011; Myers et al. 2014; Wang et al. 2017). Fractional anisotropy (FA) is a metric derived from diffusion MRI that encodes the directional preference of water diffusion and thus indirectly describes the degree of directionality of white matter pathways. FA of the dorsal pathways, including the left arcuate fasciculus, at age 6 years, were found to predict reading skills at age 8 years, even after consideration of demographic covariates (sex and socioeconomic status) and individual variation in pre-literacy skills (language abilities and phonological awareness), all of which were correlated with reading outcome (Borchers et al., 2019). Similarly, Yeatman and colleagues (2012) demonstrated that the rates of white matter development for the left arcuate fasciculus and the left inferior longitudinal fasciculus predicted 43% of the variance in reading scores in children from age 7 to 12 years. Strikingly, the pattern of white matter development in this study differed among children: above-average readers had initially lower FA that increased over time, while below-average readers had initially higher FA that decreased over time (Yeatman et al., 2012). Wang and colleagues (2017) described a similar pattern across early reading development in children with and without a familiar risk for dyslexia: FA was positively associated with reading development, and subsequent good readers showed faster white matter development in the arcuate fasciculus compared to poor readers. Taken together, these data suggest that developmental changes in white matter may play an important role in typical and atypical reading development.

Few studies to date have evaluated associations between the development of reading skills in relation to the microstructure of neural pathways outside of the language-reading network. In this study, we focused on the optic radiations (OR), circuits alternatively labelled the geniculocalcarine tracts, the geniculostriate pathways, or posterior thalamic radiations. Axons in this circuit course from the lateral geniculate nucleus to primary visual cortex in the occipital lobe. Cross-sectional data show that FA of the OR increases from age 5 to 18 years (Dayan et al. 2015). The OR may prove critical for pre-linguistic stages of reading, such as carrying visual and spatial information of print from the eye to visual cortex to begin the reading process. Supporting evidence is that visual processes have been associated with reading skills and reading disorders in a dynamic interplay over development (Yeatman and White 2021). Furthermore, the OR circuit has been implicated in reading; a subgroup of individuals with a genetic variant of dyslexia showed differences in the OR in association with deficits of visual motion processing at early stages of visual analysis (Perani et al. 2021). A recent study found correlations between microstructural properties of the OR and single word reading skills in a very large cross-sectional sample of children from ages 6 to 16 years (Koirala et al. 2021). However, it is unclear whether the development of children’s reading abilities during the learning process relates to maturational changes in OR.

In the present study, we evaluated the dynamics of reading development and the OR in two groups, children born at term (FT) and children born very preterm (PT: less than 32 weeks gestational age at birth), both with a wide range of reading abilities. Children born PT are an important comparison group for several reasons. First, they are at risk of developing poor reading skills (Aylward 2014; Bhutta et al. 2002; Kovachy et al. 2015). They learn to read but on average perform below their FT peers, and this gap has been shown to increase between ages 6 and 12 years (Kovachy et al. 2015). Second, children born PT are at risk for white matter injuries based on complications of prematurity (Volpe 2009). Diffusion MRI studies have revealed differences in microstructural properties of white matter pathways in children born PT relative to children born FT, even in the absence of significant white matter lesions on clinical MRI scans. Differences can be seen during the neonatal period (Anjari et al. 2007; Giménez et al. 2008; Rose et al. 2008) and persist to school age and beyond to adulthood (Allin et al. 2011; Dodson et al. 2017; Eikenes et al. 2011; Nagy et al. 2003). Third, microstructural properties various tracts have been found to relate to function in children born PT. Microstructural properties of the optic radiations have been associated with visual functioning in children born PT (Bassi et al. 2008). Interestingly, the associations of microstructural properties of white matter pathways and reading skills have been found to differ in children born PT and FT (Bruckert et al. 2019; Dodson et al. 2018; Frye et al. 2011; Mullen et al. 2011; Travis, Ben-Shachar, et al. 2016). For example, Frye and colleagues found that diffusion MRI metrics of the left superior longitudinal fasciculus was linked to concurrent reading skills in adolescents born PT but not in FT peers. By contrast, Travis et al (2016) found that the direction of concurrent white matter-reading associations differed between children born FT and PT with negative associations in the FT group and positive associations in the PT group (Travis et al., 2016). These findings are in a line with a recent longitudinal study that found that diffusion MRI metrics of the dorsal pathways at age 6 years were associated with reading outcome two years later in children born FT, but not in children born PT (Bruckert et al. 2019). Data linking reading to white matter pathways beyond the classic dorsal and ventral streams of the language and reading pathways are very limited in preterm samples.

The current longitudinal study had three specific aims: (1) To determine if reading skills at age 6 and at age 8 would correlate with microstructural properties of the OR, non-language-related white matter circuits that may be involved in sending visual information from regions of the brain required for the analysis of print to regions involved in processing phonological or orthographic information. (2) To evaluate whether growth in reading skills during the dynamic period from ages 6 to 8 years would be associated with changes in white matter characteristics of the optic radiations during that same epoch. (3) To assess whether reading development and properties of the OR would be similar in children born FT and PT and whether associations between growth in reading and change in the microstructure of the FA are similar in the two birth groups. This study was designed to increase our understanding of how individual differences in reading relate to microstructural properties of white matter circuits beyond language circuits and how the cumulative processes of learning and brain development interact early in the development of reading. Including children born preterm helps us determine if developmental pathways in the brain-behavior relations can vary substantially across clinical populations. We hypothesized that i) reading skills would increase over time in both groups of children, ii) FA of the OR would increase over time in both groups, iii) similar to language circuits, concurrent associations of reading and FA of the OR and associations of changes in FA of the OR and growth in reading abilities would be significant, and iii) based on previous findings (Bruckert et al. 2019; Travis, Ben-Shachar, et al. 2016; Travis et al. 2019; Dodson et al. 2018), birth group status (FT vs PT) would moderate the pattern of association between white matter change of the optic radiations and reading growth, indicating distinctive patterns of brain-behavior development in children born at term and preterm.

## 2. Methods

### 2.1 Participants

Children participated in a longitudinal study that examined the neural basis of reading and were recruited from the San Francisco Bay Area (Borchers et al., 2019; Travis et al., 2016). Children that were born at ≥37 weeks gestational age or had a birth weight ≥2,500 g were defined as FT. Children that were born at ≤ 32 weeks gestational age were defined as PT; these children are at high risk for white matter injury (Volpe, 2009) and decrements in reading ability (Aarnoudse-Moens et al., 2009; Kovachy et al., 2015). All children met the following inclusion criteria: i) no history of a neurological or medical condition (other than prematurity or its complications) that might impact learning to read such as genetic disorders, significant hearing loss or visual impairment, (ii) English speakers with at least 2 years of English exposure, and (iii) non-verbal and verbal IQ as assessed by the Wechsler Abbreviated Scale of Intelligence (WASI-II; Wechsler & Hsaio-pin, 2011) were >85. Our cohort included 34 children born FT and 34 children born PT (Table 1) who were a subset of a larger sample who have been described in previous studies (Borchers et al. 2019; Travis, Ben-Shachar, et al. 2016; Dodson et al. 2017; Bruckert et al. 2019). Participants were included in this current analysis if they had usable dMRI studies at both ages and reading assessments at three ages. We obtained informed consent from a parent or guardian for all children included in this study; all children were compensated for behavioral and MRI sessions. The research protocol was approved by the Stanford University Institutional Review Board.

**Table 1.** Demographic characteristics, reading scores, and diffusion data of the cohort (significant p-values are printed in bold).

| **N<sub>total</sub> = 68** | **Full term**<br>**(n = 34)**<br>N (%) or *M* (*SD*) | **Preterm**<br>**(n = 34)**<br>N (%) or *M* (*SD*) | ***U*<sup>\*</sup> or *X*<sup>2</sup>** | ***p*-value** |
| --- | --- | --- | --- | --- |
| <u>*Demographic characteristics*</u> |  |  |  |  |
| Male | 16 (47.1%) | 22 (64.7%) | 2.15 | .143 |
| Age at time 1 | 6.2 yrs (2.3 mon) | 6.2 yrs (2.3 mon) | -0.80 | .429 |
| Age at time 2 | 7.1 yrs (1.7 mon) | 7.2 yrs (2.4 mon) | 0.86 | .392 |
| Age at time 3 | 8.2 yrs (2.0 mon) | 8.2 yrs (2.3 mon) | 1.33 | .188 |
| SES | 58.8 (10.1) | 54.6 (12.6) | 422 | .054 |
| GA (weeks) | 39.5 (1.54) | 29.6 (2.50) | 18.5 | < **.001** |
| Birthweight (grams) | 3361 (447) | 1323 (501) | 9.00 | < **.001** |
| <u>*Reading Scores: Word Identification raw scores from the WRMT-3*</u> |  |  |  |  |
| Age 6 yrs | 12.4 (10.4) | 8.3 (8.74) | 355 | .128 |
| Age 7 yrs | 19.6 (8.02) | 16.9 (7.22) | 424 | .085 |
| Age 8 yrs | 26.2 (6.92) | 24.6 (5.06) | 455 | .130 |
| <u>*Diffusion Data: Fractional anisotropy*</u> |  |  |  |  |
| OR-R at age 6 yrs | 0.514 (0.076) | 0.508 (0.056) | 537 | .615 |
| OR-R at age 8 yrs | 0.580 (0.096) | 0.535 (0.061) | 313 | **.001** |
| OR-L at age 6 yrs | 0.514 (0.063) | 0.502 (0.064) | 519 | .469 |
| OR-L at age 8 yrs | 0.587 (0.063) | 0.545 (0.064) | 357 | **.007** |

SES, socioeconomic status from a modified version of the Hollingshead Four Factor Index of Socioeconomic Status (Hollingshead, 1975); GA, gestational age; WRMT-3, Woodcock-Johnson Reading Mastery Test, Third Edition; yrs years; mon months. \* Non-parametric Mann-Whitney U

### 2.2 Procedures

Children born FT and PT were enrolled at age 6 years and followed up at age 7 and 8 years. The assessment of demographic data and cognitive abilities has been described previously (Borchers et al. 2019; Travis, Ben-Shachar, et al. 2016) and is briefly summarized below. At age 6 years, parents completed a comprehensive demographic and health questionnaire. Socioeconomic status (SES) was measured using a modified version of the Hollingshead Four Factor Index of Socioeconomic Status (Hollingshead, 1975), which takes into account both parents education and occupation (range = 8 to 66).

Reading skills were assessed using the Woodcock Reading Mastery Test – Third Edition (WRMT-III; Woodcock, 2011) at all three visits. We used Word Identification (WID), an untimed overt reading of real words, as our main outcome measure. Raw scores, rather than age-adjusted standard scores, were used to allow analysis of change over time.

Brain MRI data were acquired at age 6 and 8 years only using a 3T scanner (GE MR750 Discovery; GE Healthcare, Waukesha, WI, USA) with a 32-channel head coil. All children were scanned for research purposes and without the use of sedation. MRI data analyzed in the current study included i) a high resolution T1-weighted scan using a 3D fast-spoiled gradient (FSPGR) sequence (TR = 7.24 ms; TE = 2.78 ms; FOV = 230 mm × 230 mm; acquisition matrix = 256 × 256; 0.9 mm isotropic voxels; orientation = sagittal) and ii) a diffusion MRI scan using a dual-spin echo, echo-planar imaging sequence (96 directions, b = 2500 s/mm^2^, 3 b =0 volumes, voxel size = 0.8549 × 0.8549 × 2 mm^3^,TR = 8300 ms, TE = 83.1 ms).

### 2.3 MRI analysis

MRI data were managed and analyzed using a neuroinformatics platform (Flywheel.io) that guarantees data provenance, implements reproducible computational methods, and facilitates data sharing. The diffusion MRI analysis pipeline consisted of three main steps: (i) defining regions of interest (ROIs) for tract identification, (ii) diffusion MRI preprocessing and (iii) whole-brain tractography and tract segmentation. The steps were described in detail by Lerma-Usabiaga et al. (2019) and Liu et al. (2022) and are briefly summarized below.

#### 2.3.1 ROI definition

The ROIs used for the identification of the OR were segmented in or transferred to the child’s anatomical T1-weighted image and included: (i) the lateral geniculate nucleus (LGN), (ii) the visual cortex (areas V1 and V2), and (iii) an additional inclusion ROI to improve neuroanatomical accuracy of the OR (Liu et al. 2022). For the left and right LGN, Freesurfer (http://surfer.nmr.mgh.harvard.edu/) was used to perform cortical and subcortical parcellation of each child’s T1-weighted image. This step included running the thalamic segmentation module implemented in Freesurfer to identify LGN bilaterally (Iglesias et al. 2018). For the visual cortex, the Neuropythy tool (Benson & Winawer 2018) was run on the Freesurfer results to perform visual cortex parcellation. The resulting V1 and V2 ROIs were combined to create the left and right visual cortex ROIs (Liu et al. 2022). To ensure that the ROIs of the LGN and visual cortex extended to the interface of grey and white matter, they were dilated by three and two cubic voxels, respectively. The additional inclusion ROI was drawn in MNI space (coronal plane of y = -80 encompassing the posterior limb of the internal capsule) and transformed to native space using non-linear registration. This was done to include OR fibers that pass through the internal capsule (Liu et al. 2022).

#### 2.3.2 Diffusion MRI preprocessing

Diffusion MRI data were preprocessed using a combination of tools from MRtrix3 (github.com/MRtrix3/mrtrix3), FSL (https://fsl.fmrib.ox.ac.uk/), and mrDiffusion (http://github.com/vistalab) as implemented in the Reproducible Tract Profiles (RTP) pipeline (Lerma-Usabiaga et al., 2019). In brief, we (i) denoised the data using principal component analysis (Veraart et al., 2016a; Veraart et al., 2016b) and Gibbs ringing correction (Kellner et al., 2016), (ii) applied FSL’s eddy current correction (Andersson and Sotiropoulos, 2016), (iii) resampled the data to 2 x 2 x 2 mm^3^ isotropic voxels, and (iv) aligned the diffusion data to the average of the non-diffusion-weighted volumes, which were aligned to the child’s high-resolution anatomical image using rigid body transformation.

#### 2.3.3 Diffusion MRI tractography

The preprocessed diffusion MRI data served as the input for whole-brain tractography and tract segmentation. We used the constrained spherical deconvolution model (CSD, Tournier et al., 2007) with eight spherical harmonics (lmax = 8) to calculate fiber orientation distributions (FOD) for each voxel. These FODs were used for diffusion MRI tractography, which consisted of the following steps: (i) Ensemble Tractography (Takemura et al., 2016) to estimate the whole-brain white matter connectome. MRtrix3 (Tournier et al., 2019) was used to generate three candidate connectomes that varied in their minimum angle parameters (50°, 30°, 10°). For each candidate connectome, a probabilistic tracking algorithm (iFOD2) was used with a step size of 1 mm, a minimum length of 10 mm, a maximum length of 200 mm, and an FOD stopping criterion of 0.04. (ii) Spherical-deconvolution Informed Filtering of Tractograms (SIFT) to improve the quantitative nature of the ensemble connectome by filtering the data such that the streamline densities match the FOD lobe integral. The resulting ensemble connectome retained 500,000 streamlines. (iii) Concatenation of the three candidate connectomes into one ensemble connectome. (iv) Automated Fiber Quantification (AFQ, Yeatman et al., 2012) to segment the resulting whole-brain connectome of each child into our two tracts of interest: left and right OR. Tract segmentation was done using a way-point ROI approach as described by Wakana et al. (2007). We used the three ROIs that were created in the first step of the pipeline to segment the left or right OR. Figure 1 shows the ROIs and resulting tracts. The tracts were refined by removing streamlines that were more than 5 standard deviations away from the core of the tract or that were more than 4 standard deviations above the mean streamline length (Yeatman et al., 2012). While CSD was used to model the fibers (as it can discern crossing fibers), the diffusion tensor model (DTI) was used to calculate FA because it is a commonly used diffusion metric that is readily interpretable and for which there are developmental data. Calculation of mean tract-FA for the left and right OR was restricted to the region between the LGN and visual cortex as defined by the corresponding ROIs.

**Figure 1.**
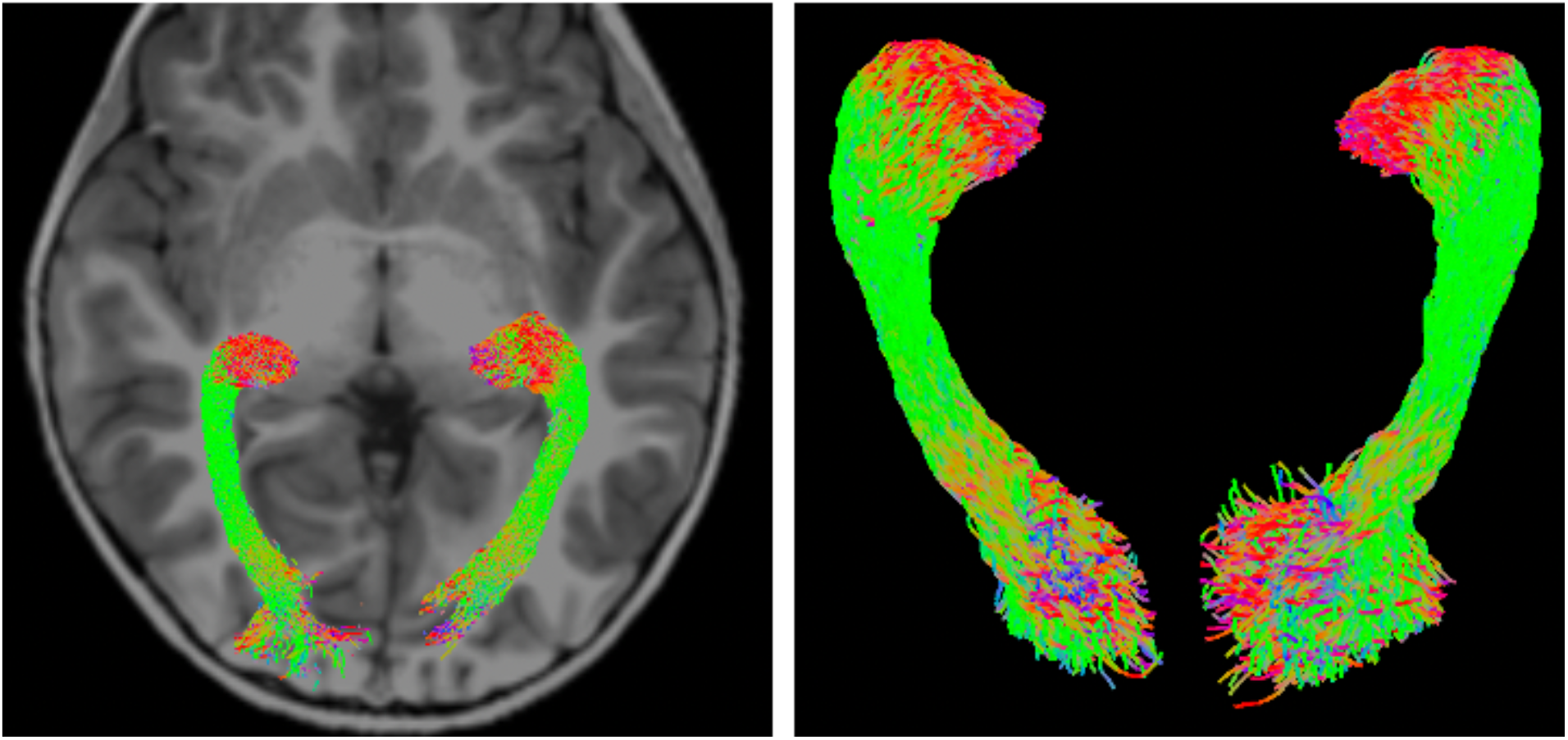
Diffusion MRI tractography of bilateral optic radiation. Left panel shows tract renderings displayed on a T1-weighted image from a representative child. Right panel shows 3D renderings of the corresponding tracts.

### 2.4 Statistical analysis

We performed all statistical analyses in SPSS (version 23.0, IBM Corporation, 2014) and R version 4.0.2 (R Core Team, 2020) with lme4 (Bates et al., 2015) and sjstats (Lüdecke, 2018). Statistical significance was set to p < 0.05. Descriptive statistics were used to characterize the sample. Chi-square tests and non-parametric Mann-Whitney U-Tests were used to examine differences between children born FT and PT on demographic variables, reading scores, and FA of the OR. Correlational analyses were used to assess association of mean tract-FA values of the left and right OR and to establish whether mean FA of the OR at age 6 and at age 8 was associated with concurrent reading at age 6 and at age 8 years. We modeled individual growth trajectories over age and explored predictors of individual differences in trajectories of linear change over time. Growth curve modeling is the preferred approach to analyzing repeated-measures data and describing developmental patterns (Boyle & Willms, 2001; Curran et al., 2010). We conducted an unconditional model examining reading growth as indexed by WID, time (age 6, 7, and 8 years) was the within-subjects factor and birth group (FT vs. PT) was the between-subjects factor both treated as fixed effects. Participant was entered as a random effect to capture variation associated with each participant’s intercept. The intercept can be set at any time point. We set the intercept at age 7 years so the fixed effect represented mean level of performance midway through the time frame. Next, we added change in FA of left or right OR from age 6 to 8 years as predictors of trajectories of growth in reading scores.

## 3. Results

### 3.1 Characteristics of the sample

Table 1 shows demographic characteristics of the children born FT and PT. The groups were matched in proportion of males, age at all three visits, and SES. Mean SES in the overall sample was 56.7 (SD = 11.5) indicating that children were generally from high SES backgrounds. By design, children born FT were significantly older in gestational age and had higher birthweights than children born PT. Raw scores on the reading measures are also shown in Table 1. While the mean reading scores were higher for the FT than PT group at age 6, 7, and 8, the differences did not achieve statistical significance.

Mean tract-FA values of the left and right OR increased between age 6 and 8 in both groups. Mean tract-FA values of the left and right OR were generally higher in the FT than the PT group. These differences achieved statistical significance at age 8 years, but not at age 6. Because mean tract-FA of the left and right OR were highly correlated at age 6 years (*rP* = 0.89, *p* < 0.001) and age 8 years (*rP* = 0.81, *p* < 0.001), we computed mean tract-FA across the two hemispheres and used the averaged bilateral OR FA values in all subsequent analyses to reduce the number of comparisons.

### 3.2 Reading scores and growth in reading from age 6 to 8 years

Table 2, Model 1A shows the linear mixed-effects model of reading growth from ages 6 to 8 years for children born FT and PT. The fixed effect of time was significant, whereas the fixed effect of group and the group-by-time interaction did not achieve statistical significance. These results are depicted in Figure 2. These analyses show that both children born FT and PT demonstrated increasing reading scores over time, from age 6 to 8 years, indicating reading growth.

**Table 2.** Estimated models of reading growth trajectory from ages 6 to 8 years for children born full term and preterm before and after adding FA change of the optic radiation (averaged across left and right hemispheres; OR).

| OR | Model 1A | Model 1B | Model 1C | Model 1D |
| --- | --- | --- | --- | --- |
| Time | **6.91 (0.48)** *** | **6.91 (0.48)** *** | **6.91 (0.48)** *** | **6.85 (0.55)** *** |
| Group | -2.83 (1.70)<sup>+</sup> | **-3.48 (1.67)**<sup>*</sup> | **-5.00 (1.85)** ** | **-3.48 (1.67)**<sup>*</sup> |
| Time x Group | 1.23 (0.71)<sup>+</sup> | 1.23 (0.71)<sup>+</sup> | 1.23 (0.71)<sup>+</sup> | 1.26 (0.72)<sup>+</sup> |
| Δ OR | - | **-19.3 (9.00)**<sup>*</sup> | **-29.4 (10.5)** ** | **-19.3 (9.00)**<sup>*</sup> |
| Δ OR x Group | - | - | 33.4 (19.2)<sup>+</sup> | - |
| Δ OR x Time | - | - | - | 0.83 (3.86) |
| R<sup>2</sup> (fixed effects) | 39% | 42% | 45% | 42% |
| Log Likelihood | -623.0 | -620.8 | -619.3 | -620.8 |

Data are estimates (SE) of the fixed effects. Significant values are printed in bold. <sup>+</sup>p < 0.10, <sup>\*</sup>p < 0.05, <sup>\*\*</sup>p < 0.01, <sup>\*\*\*</sup>p < 0.001. r<sup>2</sup>-change for Model 1B is in relation to Model 1A; r<sup>2</sup>-change for Models 1C and 1D are in relation to Model 1B.

**Figure 2.**
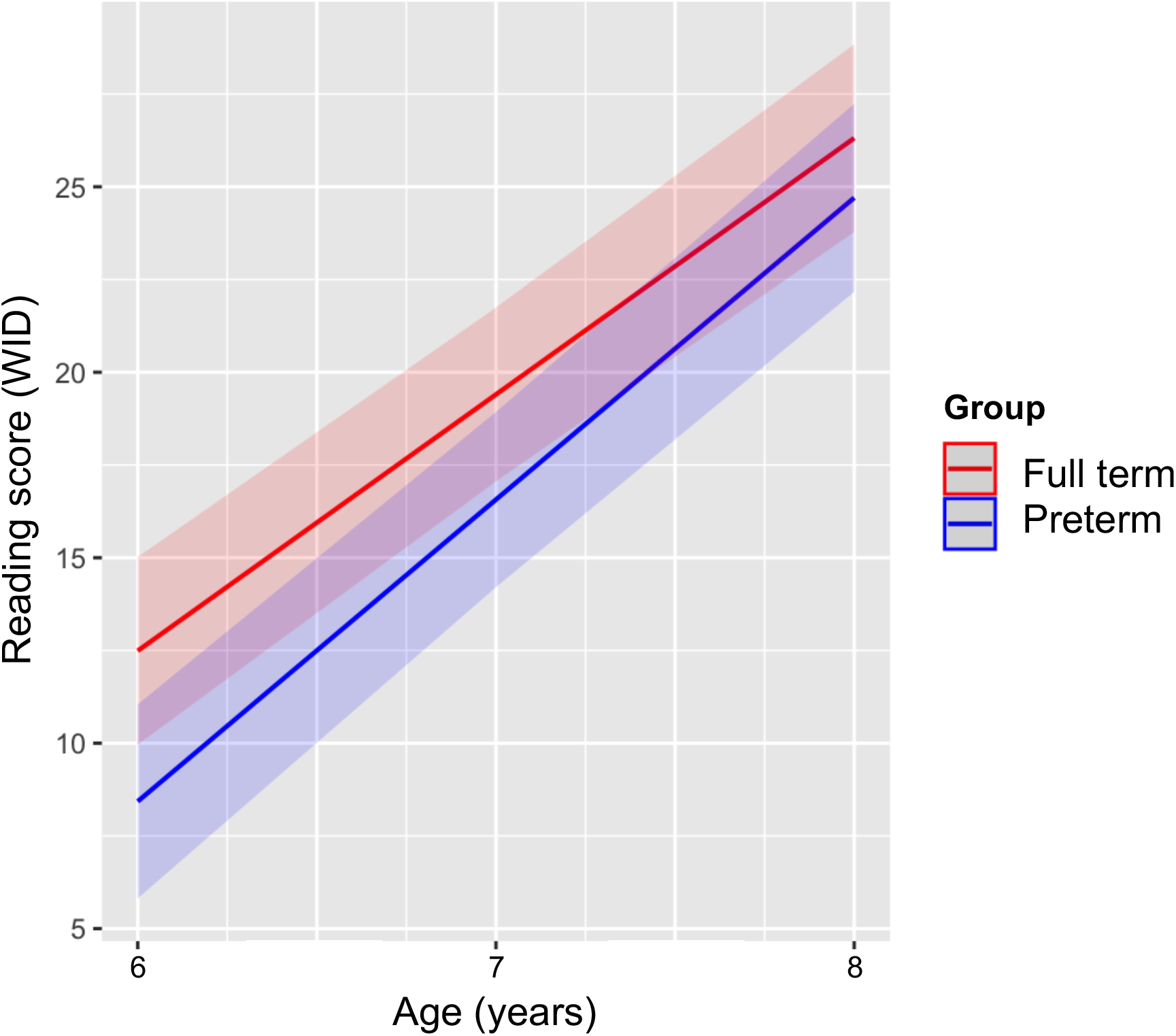
Predicted scores of word identification (WID) from the Woodcock Reading Mastery Test for children born full term (red) and preterm (blue).

### 3.3 Average bilateral OR FA in relation to reading scores

Average bilateral OR FA was weakly associated with reading scores at age 6 years (*r* = 0.27, *p* = 0.037) but was not associated at age 8 years (*r* = -0.07, *p* = 0.573).

Table 2, Model 1B, shows that the main effect of change in average bilateral OR FA (FA change) significantly increased the overall model fit, added 3% unique variance. Figure 3 plots this model, demonstrating the effect of FA change on reading growth. Models 1C and 1D show that adding the group-by-tract and group-by-time interactions did not significantly increase overall model fit. Overall, these results indicate that in both the FT and PT groups, the growth of reading was associated with the degree of change in FA of the OR. ThIS association was negative; faster growth rate in reading skills was associated with decreasing FA

**Figure 3.**
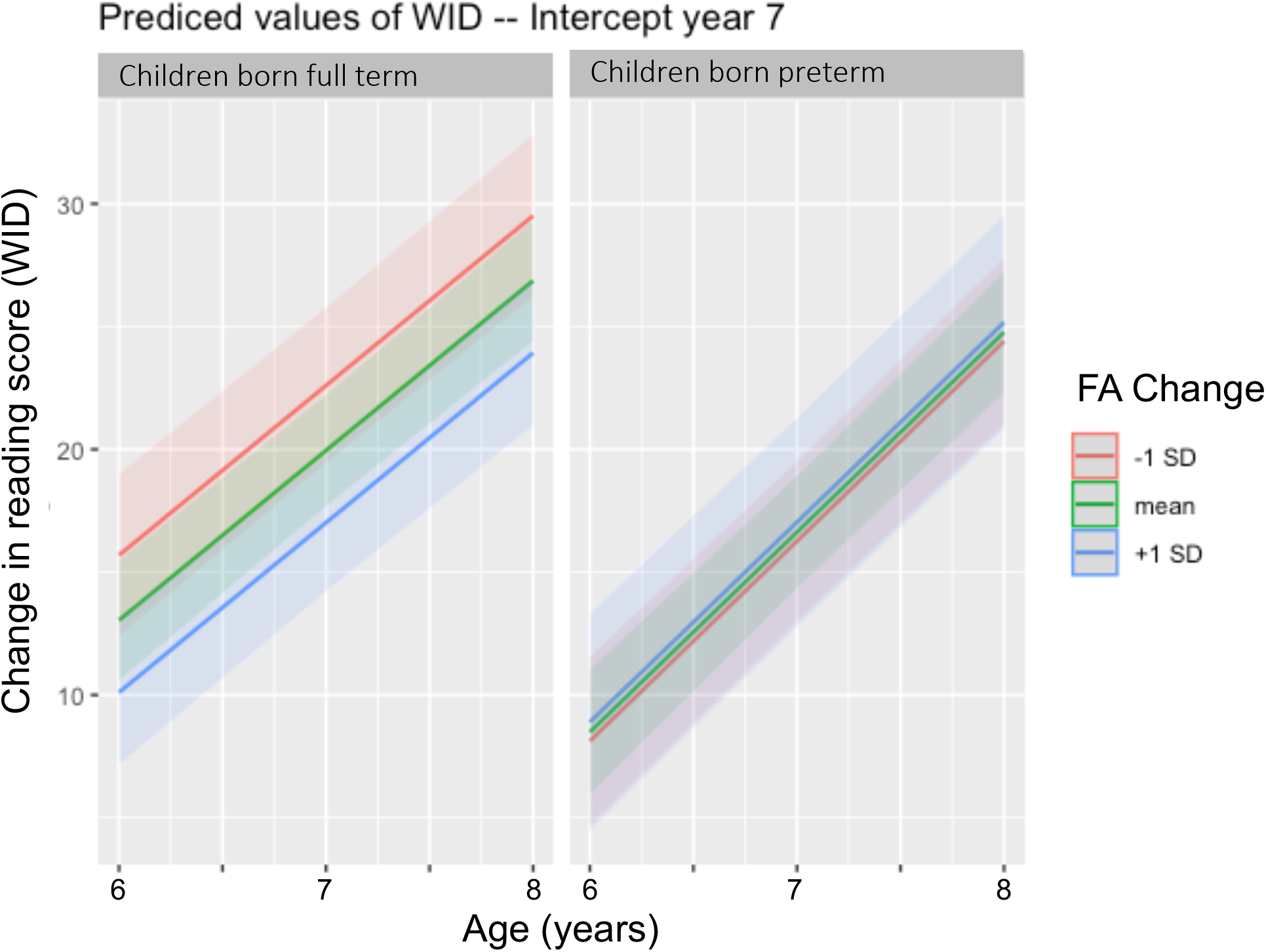
Modeled word identification (WID) scores from the Woodcock Reading Mastery Test for children born full term (left panel) and preterm (right panel) as a function of change in average bilateral optic radiations fractional anisotropy (FA).

## 4. Discussion

This study increased our understanding of how individual differences in reading relate to microstructural properties of white matter circuits beyond the left-lateralized language network. We analyzed the optic radiations, reasoning that a visual pathway might be relevant to reading development because reading begins with visual analysis of print. The longitudinal analyses demonstrated how the cumulative processes of learning and brain development interact early in the development of reading. The inclusion of a clinical group, children born preterm, was designed to determine if developmental pathways in the brain-behavior relations can vary substantially across clinical populations.

As anticipated, reading skills increased over time in both birth groups. Despite previous data showing poorer reading skills in children born PT than FT (Aylward 2014; Bhutta et al. 2002; Kovachy et al. 2015), in this sample, rates of growth were comparable in the FT and PT groups. We attribute lack of birth group differences to our small sample size and the likely educational advantages of this high SES PT cohort.

A novelty of the current study was analyzing microstructure of the OR in relation to reading. One obstacle has been accuracy of reconstructing the OR from deterministic tractography (Lilja et al. 2014; Winston 2013). For this study, we used probabilistic tractography in the child’s native space and native-space segmentation of the thalamus and visual cortex. This method has proven useful in a different clinical population (Bruckert et al, under review) and gives consistent and robust tracking results (Figure 1). The results of FA of the OR in this study are further validation of the methods. We learned that FA of the OR increased between ages 6 and 8 years in both birth groups. These findings are consistent with previous studies that show increasing FA of other tracts with age in healthy FT children (Yeatman, Wandell, and Mezer 2014; Catherine Lebel and Beaulieu 2011) and in children born PT (Schneider et al. 2016). Increases in FA with age may indicate several microstructural changes in the neurobiological of white matter, including increased myelination and greater compactness of fibers, both of which would contribute to increasing FA. Moreover, FA was higher in the FT than PT groups and the difference reached statistical significance when the children reached age 8 years. Birth group difference in FA of white matter circuits have been found in previous studies, though the direction of the difference has been found to vary as a function of the specific tract analyzed (Dodson et al. 2017; Travis et al. 2015).

The results showed that concurrent correlations of average bilateral OR FA and reading were weakly significant at age 6, suggesting that transfer of visual information is modestly associated with individual differences in very early stages of reading. However, concurrent correlations became insignificant at age 8. One hypothesis based on these results is that as children become competent readers, visual processing and/or the transfer of visual-spatial information to orthographic or phonological brain centers becomes sufficiently efficient that individual differences in the visual circuit are no longer contributory to individual differences in reading. Future studies could determine if the concurrent associations of FA of the OR and reading varies as a function of reading abilities rather than age.

The key findings of this study were that reading growth from age 6 to 8 years was associated with change in FA of the OR and that the direction of the association was negative. In simple terms, greater decreases in average FA of the OR FA were associated with more rapid reading growth. Negative associations of FA and reading have been previously reported at a single point in time ((Bruckert et al. 2020), across ages in a cross-sectional study (Travis, Ben-Shachar, et al. 2016), and across ages in a longitudinal study (Yeatman et al. 2012). In this study, we found a negative association between reading development and FA change even though both children born FT and PT showed increases in FA between ages 6 and 8 years. These findings are strong evidence that the dynamic factors associated with the development of white matter pathways are not necessarily the same dynamic factors associated with individual differences in the development. For example, the development of white matter pathways may rely on increased myelination and axonal packing, resulting in increased FA. However, the development of reading may rely on increased axonal diameter or complexity of fiber bundles. We must conceptualize white matter development as a interplay of multiple underlying neurobiological factors, each of which may contribute differentially to functional outcomes. On this basis, the future of white matter studies of brain-behavior relations should consider use of multiple imaging methods, each sensitive to different underlying factors. The integration of the results will be more revealing than the results of any single imaging method in isolation.

We hypothesized that the pattern of association between FA change and reading growth would differ as a function of birth group. In the current study, the interaction between FA-change and birth group did not achieve statistical significance. Previous studies have shown that the pattern of associations vary between children born FT and PT (Travis et al., Lisa prediction), By contrast, the findings here suggest that the dynamic factors associated with white matter development in children born PT may not differ significantly from the factors associated with white matter development in children born FT. It could be that in the OR and the mixed model analyses, our modest sample size was not adequate to detect an interaction effect. Further exploration of birth group differences should include a larger longitudinal samples and the use of multiple MRI methods that are sensitive to the different properties of white development.

## 5. Conclusion

This study suggests that the white matter circuitry underlying the development of reading extends beyond language circuitry and includes the ORs. Dynamic change in the OR over time, not just the microstructure of OR at a single age, contribute to the variance in reading development. The direction of associations suggests that individual differences in reading development are related to microstructural features of white matter that are distinct from the features associated with white matter development. In this study, the patterns of association of development of reading and change in FA of the OR were comparable in children born FT and PT.

## Data Availability

All data produced in the present study are available upon reasonable request to the authors.

## References

Allin, Matthew P. G., Dimitris Kontis, Muriel Walshe, John Wyatt, Gareth J. Barker, Richard A. A. Kanaan, Philip McGuire, Larry Rifkin, Robin M. Murray, and Chiara Nosarti. 2011. “White Matter and Cognition in Adults Who Were Born Preterm.” Edited by Joseph Najbauer. PLoS ONE 6 (10): e24525. https://doi.org/10.1371/journal.pone.0024525.

Anjari, Mustafa, Latha Srinivasan, Joanna M. Allsop, Joseph V. Hajnal, Mary A. Rutherford, A. David Edwards, and Serena J. Counsell. 2007. “Diffusion Tensor Imaging with Tract-Based Spatial Statistics Reveals Local White Matter Abnormalities in Preterm Infants.” NeuroImage 35 (3): 1021–27. https://doi.org/10.1016/J.NEUROIMAGE.2007.01.035.

Aylward, Glen P. 2014. “Neurodevelopmental Outcomes of Infants Born Prematurely.” Journal of Developmental & Behavioral Pediatrics 35 (6): 394–407. https://doi.org/10.1097/01.DBP.0000452240.39511.d4.

Bassi, Laura, Daniela Ricci, Anna Volzone, Joanna M. Allsop, Latha Srinivasan, Aakash Pai, Carmen Ribes, et al. 2008. “Probabilistic Diffusion Tractography of the Optic Radiations and Visual Function in Preterm Infants at Term Equivalent Age.” Brain 131 (2): 573–82. https://doi.org/10.1093/BRAIN/AWM327.

Beaulieu, Christian, Christopher Plewes, Lori Anne Paulson, Dawne Roy, Lindsay Snook, Luis Concha, and Linda Phillips. 2005. “Imaging Brain Connectivity in Children with Diverse Reading Ability.” NeuroImage 25 (4): 1266–71. https://doi.org/10.1016/j.neuroimage.2004.12.053.

Ben-Shachar, Michal, Robert F Dougherty, and Brian A Wandell. 2007. “White Matter Pathways in Reading.” Current Opinion in Neurobiology 17 (2): 258–70. https://doi.org/10.1016/j.conb.2007.03.006.

Benson, Noah C., and Jonathan Winawer. 2018. “Bayesian Analysis of Retinotopic Maps.” ELife 7 (December). https://doi.org/10.7554/ELIFE.40224.

Bhutta, Adnan T., Mario A. Cleves, Patrick H. Casey, Mary M. Cradock, and K. J.S. Anand. 2002. “Cognitive and Behavioral Outcomes of School-Aged Children Who Were Born Preterm: A Meta-Analysis.” JAMA 288 (6): 728–37. https://doi.org/10.1001/JAMA.288.6.728.

Borchers, Lauren R, Lisa Bruckert, Cory K Dodson, Katherine E Travis, Virginia A Marchman, Michal Ben-Shachar, and Heidi M Feldman. 2018. “Microstructural Properties of White Matter Pathways in Relation to Subsequent Reading Abilities in Children: A Longitudinal Analysis.” *Brain Structure and Function*, December, 1–15. https://doi.org/10.1007/s00429-018-1813-z.

Borchers, Lauren R, Lisa Bruckert, Katherine E Travis, Cory K Dodson, Irene M Loe, Virginia A Marchman, and Heidi M Feldman. 2019. “Predicting Text Reading Skills at Age 8 Years in Children Born Preterm and at Term.” Early Human Development 130 (March): 80–86. https://doi.org/10.1016/J.EARLHUMDEV.2019.01.012.

Bruckert, Lisa, Lauren R Borchers, Cory K Dodson, Virginia A Marchman, Katherine E Travis, Michal Ben-Shachar, and Heidi M Feldman. 2019. “White Matter Plasticity in Reading-Related Pathways Differs in Children Born Preterm and at Term: A Longitudinal Analysis.” Frontiers in Human Neuroscience 13 (May): 139. https://doi.org/10.3389/fnhum.2019.00139.

Bruckert, Lisa, Katherine E. Travis, Aviv A. Mezer, Michal Ben-Shachar, and Heidi M. Feldman. 2020. “Associations of Reading Efficiency with White Matter Properties of the Cerebellar Peduncles in Children.” Cerebellum 19 (6): 771–77. https://doi.org/10.1007/s12311-020-01162-2.

Darki, Fahimeh, Myriam Peyrard-Janvid, Hans Matsson, Juha Kere, and Torkel Klingberg. 2012. “Three Dyslexia Susceptibility Genes, DYX1C1, DCDC2, and KIAA0319, Affect Temporo-Parietal White Matter Structure.” Biological Psychiatry 72 (8): 671–76. https://doi.org/10.1016/J.BIOPSYCH.2012.05.008.

Dayan, Michael, Monica Munoz, Sebastian Jentschke, Martin J. Chadwick, Janine M. Cooper, Kate Riney, Faraneh Vargha-Khadem, and Chris A. Clark. 2015. “Optic Radiation Structure and Anatomy in the Normally Developing Brain Determined Using Diffusion MRI and Tractography.” Brain Structure & Function 220 (1): 291–306. https://doi.org/10.1007/S00429-013-0655-Y.

Deutsch, Gayle K, Robert F Dougherty, Roland Bammer, Wai Ting Siok, John D E Gabrieli, and Brian A Wandell. 2005. “Children’s Reading Performance Is Correlated with White Matter Structure Measured by Diffusion Tensor Imaging.” Cortex 41 (3): 354–63. https://doi.org/10.1016/S0010-9452(08)70272-7.

Dodson, Cory K, Katherine E Travis, Michal Ben-Shachar, and Heidi M Feldman. 2017. “White Matter Microstructure of 6-Year Old Children Born Preterm and Full Term.” NeuroImage: Clinical 16: 268–75. https://doi.org/10.1016/j.nicl.2017.08.005.

Dodson, Cory K, Katherine E Travis, Lauren R Borchers, Virginia A Marchman, Michal Ben-Shachar, and Heidi M Feldman. 2018. “White Matter Properties Associated with Pre-Reading Skills in 6-Year-Old Children Born Preterm and at Term.” Developmental Medicine & Child Neurology 60 (7): 695–702. https://doi.org/10.1111/dmcn.13783.

Eikenes, Live, Gro C. Løhaugen, Ann-Mari Brubakk, Jon Skranes, and Asta K. Håberg. 2011. “Young Adults Born Preterm with Very Low Birth Weight Demonstrate Widespread White Matter Alterations on Brain DTI.” NeuroImage 54 (3): 1774–85. https://doi.org/10.1016/J.NEUROIMAGE.2010.10.037.

Frye, Richard E., Jacqueline Liederman, Khader M. Hasan, Alexis Lincoln, Benjamin Malmberg, John McLean, and Andrew Papanicolaou. 2011. “Diffusion Tensor Quantification of the Relations between Microstructural and Macrostructural Indices of White Matter and Reading.” Human Brain Mapping 32 (8): 1220–35. https://doi.org/10.1002/hbm.21103.

Giménez, Mónica, Maria J. Miranda, A. Peter Born, Zoltan Nagy, Egill Rostrup, and Terry L. Jernigan. 2008. “Accelerated Cerebral White Matter Development in Preterm Infants: A Voxel-Based Morphometry Study with Diffusion Tensor MR Imaging.” NeuroImage 41 (3): 728–34. https://doi.org/10.1016/J.NEUROIMAGE.2008.02.029.

Hickok, Gregory, and David Poeppel. 2004. “Dorsal and Ventral Streams: A Framework for Understanding Aspects of the Functional Anatomy of Language.” Cognition 92 (1–2): 67–99. https://doi.org/10.1016/j.cognition.2003.10.011.

Hoeft, Fumiko, Bruce D McCandliss, Jessica M Black, Alexander Gantman, Nahal Zakerani, Charles Hulme, Heikki Lyytinen, et al. 2011. “Neural Systems Predicting Long-Term Outcome in Dyslexia.” Proceedings of the National Academy of Sciences of the United States of America 108 (1): 361–66. https://doi.org/10.1073/pnas.1008950108.

Iglesias, Juan Eugenio, Ricardo Insausti, Garikoitz Lerma-Usabiaga, Martina Bocchetta, Koen Van Leemput, Douglas N. Greve, Andre van der Kouwe, Bruce Fischl, César Caballero-Gaudes, and Pedro M. Paz-Alonso. 2018. “A Probabilistic Atlas of the Human Thalamic Nuclei Combining Ex Vivo MRI and Histology.” NeuroImage 183 (December): 314–26. https://doi.org/10.1016/J.NEUROIMAGE.2018.08.012.

Klingberg, Torkel, Maj Hedehus, Elise Temple, Talya Salz, John D.E Gabrieli, Michael E Moseley, and Russell A Poldrack. 2000. “Microstructure of Temporo-Parietal White Matter as a Basis for Reading Ability.” Neuron 25 (2): 493–500. https://doi.org/10.1016/S0896-6273(00)80911-3.

Koirala, Nabin, Meaghan V. Perdue, Xing Su, Elena L. Grigorenko, and Nicole Landi. 2021. “Neurite Density and Arborization Is Associated with Reading Skill and Phonological Processing in Children.” NeuroImage 241 (November): 118426. https://doi.org/10.1016/J.NEUROIMAGE.2021.118426.

Kovachy, Vanessa N, Jenna N Adams, John S Tamaresis, and Heidi M Feldman. 2015. “Reading Abilities in School-Aged Preterm Children: A Review and Meta-Analysis.” Developmental Medicine and Child Neurology 57 (5): 410–19. https://doi.org/10.1111/dmcn.12652.

Lebel, C., L. Walker, A. Leemans, L. Phillips, and C. Beaulieu. 2008. “Microstructural Maturation of the Human Brain from Childhood to Adulthood.” NeuroImage 40 (3): 1044–55. https://doi.org/10.1016/J.NEUROIMAGE.2007.12.053.

Lebel, Catherine, and Christian Beaulieu. 2011. “Longitudinal Development of Human Brain Wiring Continues from Childhood into Adulthood.” Journal of Neuroscience 31 (30): 10937–47. https://doi.org/10.1523/JNEUROSCI.5302-10.2011.

Lilja, Ylva, Maria Ljungberg, Göran Starck, Kristina Malmgren, Bertil Rydenhag, and Daniel T Nilsson. 2014. “Visualizing Meyer’s Loop: A Comparison of Deterministic and Probabilistic Tractography.” Epilepsy Research 108 (3): 481–90. https://doi.org/https://doi.org/10.1016/j.eplepsyres.2014.01.017.

Liu, Mengxing, Garikoitz Lerma-Usabiaga, Francisco Clascá, and Pedro M. Paz-Alonso. 2022. “Reproducible Protocol to Obtain and Measure First-Order Relay Human Thalamic White-Matter Tracts.” NeuroImage 262 (November): 119558. https://doi.org/10.1016/J.NEUROIMAGE.2022.119558.

Mullen, Katherine M., Betty R. Vohr, Karol H. Katz, Karen C. Schneider, Cheryl Lacadie, Michelle Hampson, Robert W. Makuch, Allan L. Reiss, R. Todd Constable, and Laura R. Ment. 2011. “Preterm Birth Results in Alterations in Neural Connectivity at Age 16 Years.” NeuroImage 54 (4): 2563–70. https://doi.org/10.1016/J.NEUROIMAGE.2010.11.019.

Myers, Chelsea A., Maaike Vandermosten, Emily A. Farris, Roeland Hancock, Paul Gimenez, Jessica M. Black, Brandi Casto, et al. 2014a. “White Matter Morphometric Changes Uniquely Predict Children’s Reading Acquisition.” Psychological Science 25 (10): 1870–83. https://doi.org/10.1177/0956797614544511.

Nagy, Zoltan, Helena Westerberg, Stefan Skare, Jesper L Andersson, Anders Lilja, Olof Flodmark, Elisabeth Fernell, et al. 2003. “Preterm Children Have Disturbances of White Matter at 11 Years of Age as Shown by Diffusion Tensor Imaging.” Pediatric Research 54 (5): 672–79. https://doi.org/10.1203/01.PDR.0000084083.71422.16.

Niogi, Sumit N., and Bruce D. McCandliss. 2006. “Left Lateralized White Matter Microstructure Accounts for Individual Differences in Reading Ability and Disability.” Neuropsychologia 44 (11): 2178–88. https://doi.org/10.1016/j.neuropsychologia.2006.01.011.

Perani, Daniela, Paola Scifo, Guido M. Cicchini, Pasquale Della Rosa, Chiara Banfi, Sara Mascheretti, Andrea Falini, Cecilia Marino, and Maria Concetta Morrone. 2021. “White Matter Deficits Correlate with Visual Motion Perception Impairments in Dyslexic Carriers of the DCDC2 Genetic Risk Variant.” Experimental Brain Research 239 (9): 2725–40. https://doi.org/10.1007/S00221-021-06137-1/FIGURES/8.

Rollans, Claire, Kulpreet Cheema, George K. Georgiou, and Jacqueline Cummine. 2017. “Pathways of the Inferior Frontal Occipital Fasciculus in Overt Speech and Reading.” Neuroscience 364 (November): 93–106. https://doi.org/10.1016/J.NEUROSCIENCE.2017.09.011.

Rose, Stephen E., Xanthy Hatzigeorgiou, Mark W. Strudwick, Gail Durbridge, Peter S.W. Davies, and Paul B. Colditz. 2008. “Altered White Matter Diffusion Anisotropy in Normal and Preterm Infants at Term-Equivalent Age.” Magnetic Resonance in Medicine 60 (4): 761–67. https://doi.org/10.1002/mrm.21689.

Schneider, J, T Kober, M Bickle Graz, R Meuli, P S Hüppi, P Hagmann, and A C Truttmann. 2016. “Evolution of T1 Relaxation, ADC, and Fractional Anisotropy during Early Brain Maturation: A Serial Imaging Study on Preterm Infants.” AJNR. American Journal of Neuroradiology 37 (1): 155–62. https://doi.org/10.3174/ajnr.A4510.

Scott, Sophie K, and Richard J.S. Wise. 2004. “The Functional Neuroanatomy of Prelexical Processing in Speech Perception.” Cognition 92 (1–2): 13–45. https://doi.org/10.1016/j.cognition.2002.12.002.

Travis, Katherine E., Maria R.H. Castro, Shai Berman, Cory K Dodson, Aviv A Mezer, Michal Ben-Shachar, and Heidi M. Feldman. 2019. “More than Myelin: Probing White Matter Differences in Prematurity with Quantitative T1 and Diffusion MRI.” NeuroImage: Clinical 22: 101756. https://doi.org/10.1016/j.nicl.2019.101756.

Travis, Katherine E, Jenna N Adams, Michal Ben-Shachar, and Heidi M Feldman. 2015. “Decreased and Increased Anisotropy along Major Cerebral White Matter Tracts in Preterm Children and Adolescents.” Edited by Gaolang Gong. PLOS ONE 10 (11): e0142860. https://doi.org/10.1371/journal.pone.0142860.

Travis, Katherine E, Jenna N Adams, Vanessa N Kovachy, Michal Ben-Shachar, and Heidi M Feldman. 2016. “White Matter Properties Differ in 6-Year Old Readers and Pre-Readers.” Brain Structure and Function, September, 1–19. https://doi.org/10.1007/s00429-016-1302-1.

Travis, Katherine E, Michal Ben-Shachar, Nathaniel J Myall, and Heidi M Feldman. 2016. “Variations in the Neurobiology of Reading in Children and Adolescents Born Full Term and Preterm.” NeuroImage: Clinical 11: 555–65. https://doi.org/10.1016/j.nicl.2016.04.003.

Vandermosten, Maaike, Bart Boets, Jan Wouters, and Pol Ghesquière. 2012. “A Qualitative and Quantitative Review of Diffusion Tensor Imaging Studies in Reading and Dyslexia.” Neuroscience & Biobehavioral Reviews 36 (6): 1532–52. https://doi.org/10.1016/j.neubiorev.2012.04.002.

Volpe, Joseph J. 2009. “Brain Injury in Premature Infants: A Complex Amalgam of Destructive and Developmental Disturbances.” The Lancet. Neurology 8 (1): 110–24. https://doi.org/10.1016/S1474-4422(08)70294-1.

Wandell, Brian A, and Jason D Yeatman. 2013. “Biological Development of Reading Circuits.” Current Opinion in Neurobiology 23 (2): 261–68. https://doi.org/10.1016/j.conb.2012.12.005.

Wang, Yingying, Meaghan V. Mauer, Talia Raney, Barbara Peysakhovich, Bryce L. C. Becker, Danielle D. Sliva, and Nadine Gaab. 2017. “Development of Tract-Specific White Matter Pathways During Early Reading Development in At-Risk Children and Typical Controls.” Cerebral Cortex 27 (4): bhw095. https://doi.org/10.1093/cercor/bhw095.

Winston, Gavin P. 2013. “Epilepsy Surgery, Vision, and Driving: What Has Surgery Taught Us and Could Modern Imaging Reduce the Risk of Visual Deficits?” Epilepsia 54 (11): 1877–88. https://doi.org/https://doi.org/10.1111/epi.12372.

“WRMT-III Woodcock Reading Mastery Tests Third Edition.” n.d. Accessed September 16, 2022. https://www.pearsonassessments.com/store/usassessments/en/Store/Professional-Assessments/Academic-Learning/Reading/Woodcock-Reading-Mastery-Tests-%7C-Third-Edition/p/100000264.html?gclid=Cj0KCQjwvZCZBhCiARIsAPXbajsQDtaVV3Stj3sCkjraRtz5PmzjeEOU1oPUr-GK1ajvLMKDXYvXgCoaAqWHEALw_wcB.

Yeatman, Jason D., Brian A. Wandell, and Aviv A. Mezer. 2014. “Lifespan Maturation and Degeneration of Human Brain White Matter.” Nature Communications 5 (1): 4932. https://doi.org/10.1038/ncomms5932.

Yeatman, Jason D., and Alex L. White. 2021. “Reading: The Confluence of Vision and Language.” Https://Doi.Org/10.1146/Annurev-Vision-093019-1135097 (September): 487–517. https://doi.org/10.1146/ANNUREV-VISION-093019-113509.

Yeatman, Jason D, Robert F Dougherty, Michal Ben-Shachar, and Brian A Wandell. 2012. “Development of White Matter and Reading Skills.” Proceedings of the National Academy of Sciences of the United States of America 109 (44): E3045–53. https://doi.org/10.1073/pnas.1206792109.

